# Phenol as a breath marker for hemodialysis of chronic kidney disease patients

**DOI:** 10.1101/2024.09.02.24310987

**Authors:** Isabell Eickel, Anne-Christine Zygmunt, Frank Streit, Björn Tampe, Nils Kunze-Szikszay, Thorsten Perl

## Abstract

**Background:** We aimed to identify biomarkers in breath analysis with MCC-IMS to monitor the haemodialysis for CKD patients fast and non-invasive.

**Material & methods:** Six patients’ breath was analysed via MCC-IMS before and after dialysis and compared to blood plasma samples analysed via UPLC-FLD for potential renal failure biomarkers. Additionally, breath from six healthy control persons was analysed.

**Results:** Phenol was found as a breath marker for CKD. For three patients the phenol concentration in breath and plasma was elevated before and decreased during dialysis and reached values in the range of healthy control persons.

**Conclusion:** This study shows that the measurement of phenol via breath analysis could be used to monitor the haemodialysis for CKD-patients and might also be usable for the calculation of haemodialysis dose in the future.

## Introduction

Chronic kidney disease is a progressive disease and in end-stage renal failure patients are in need of renal transplantation or dialysis. Even though dialysis is essential for survival of patients until transplantation is possible, mortality of patients under dialysis by far exceeds mortality of a matched population without need of dialysis. One reason for this observation is the fact that current dialysis techniques are not capable of replacement of native renal function. The individualization of dialysis treatment is receiving increasing attention in research as patients with chronic kidney disease may benefit from personalized dosing of dialysis.

Renal impairment leads to accumulation of several uremic solutes including nitrogenous waste products and low molecular weight organic compounds. To prevent or treat uremic conditions dialysis is applied, causing removal of uremic toxins and water. To achieve an acceptable removal rate, a minimum dose for every treatment is necessary. Dose calculation is traditionally estimated and calculated from blood urea concentrations, dialysis treatment time and urea distribution volume (Kt/V) (Daugirdas, 2015). However, this calculation is based only on one uremic toxin, urea, with the need of invasive sampling of blood for determination baseline urea and also the disregard for other organic compounds that cumulate in renal failure.

Lipophilic, protein-bound compounds such as p-cresol or phenol for example, have been poorly represented as dialysis indicator and in evaluation of dialysis efficiency. The endogenic source of these aromatic compounds is intestinal microbacteria (Saito et al., 2018), related to tyrosine and phenylalanine metabolism. After intestinal resorption these compounds underly renal excretion. For patients with chronic renal impairment and intermittent hemodialysis (HD) an accumulation of these substances is observed (Fukuuchi et al., 2002; Gryp et al., 2017; Hida et al., 1996). An accumulation of these substances is associated with several pathologies in context of renal failure. For example, cytotoxic effects with worsening of renal function (Brocca et al., 2013; Liu et al., 2018), impaired cardiac function and coronary artery disease (Meijers et al., 2010) are described. Meanwhile, the clearance of aromatic toxins by dialysis is poor due to high protein binding and consecutively high volume of distribution (Martinez et al., 2005).

Breath gas analysis is capable of non-invasive determination and quantification of volatile substances and was demonstrated with MCC-IMS for medications like propofol (Maurer et al., 2017; Perl et al., 2009). For compounds associated with renal failure it is assumable that the volatile compounds appear in breath gas as well, as foetor uraemicus is a known finding in renal failure.

Several volatile organic compounds (VOCs) have been described as accumulated in patients with poor kidney function such as phenol, p-cresol and more (Mochalski et al., 2014). These compounds are small in size, highly volatile and low in concentration. VOCs, soluble in blood, pass the alveolar membrane and are exhaled with breath.

Ion mobility spectrometry (IMS) in combination with a multicapillary column (MCC) is a suitable measure to detect and quantify traces of compounds in complex gas mixtures. MCC– IMS may have potential medical applications in drug monitoring (Perl et al., 2009), detection of infectious diseases (Drees et al., 2019; Kunze-Szikszay et al., 2021) and further diagnostic procedures (Westhoff et al., 2009). IMS provides high sensitivity (detection limits in the ng/l, pg/l, ppb, and ppt ranges) combined with high-speed data acquisition. Due to the relatively low technical expenditure, the equipment is transportable, non-bulky, and inexpensive.

As a non-invasive method, breath analysis would be a fast and easy-to-use option for monitoring dialysis in patients needing RRT multiple times a week with sampling of exhaled breath (Vautz et al., 2010).

Breath analysis could therefore yield more steady and continuous results. It may therefore improve initial evaluation of the necessity of RRT in patients and also enable the clinicians to monitor therapy more closely and critically assess the used materials, such as filter membranes. Additionally, breath analysis may provide an opportunity to predict severe consequences of kidney failure, such as cardiac events, and chance to tackle these issues in a more timely manner.

This study was conducted as a proof of principle study to evaluate the feasibility of MCC-IMS to detect uremic volatile compounds in breath of CKD patients. Breath gas measurements with MCC-IMS were used for real-time monitoring of dialysis.

## Material and Methods

A custom-designed IMS with a β-radiation source (63Ni) for ionization was used for breath analyses. As the operating principle of MCC-IMS has been described before, only a brief description will be given here (Ruzsanyi et al., 2005). The gas of an expired air sample is preseparated by a multicapillary chromatographic column. VOC enter the drift tube with different retention times as an effect of preseparation. The gas-phase of VOCs is ionized by β-radiation. It is then injected into a drift tube where it is simultaneously attracted by an electrical field to a detector and decelerated by a counter-current gas (i.e. drift gas). The time from injection to detection describes the drift time and depends on shape and size of the compound.

The measurement program had a runtime of 16 minutes and 39 seconds. Synthetic air (scientific quality, AIR LIQUIDE Deutschland GmbH) was used as operating gas for the system. The gas flow on the multi-capillary column was set to 150 ml/min, the IMS drift gas flow was set to 100 ml/min. All experimental parameters of the MCC-IMS are summed up in **table 1**. The substances of interest were creatinine (Walser, 1998), 3-(furyl)propionic acid (Duranton et al., 2012), DL-homocysteine, L-homocystein (van Guldener et al., 2001), indoxy sulfate potassium salt (Ahmed et al., 2022), myo inositol (Melmed et al., 1977), p-cresol (Brocca et al., 2013), p-cresyl sulfate (Gryp et al., 2017), phenol (Coan et al., 1982), phenylacetic acid (Jankowski et al., 2003), sodium cyanate (Dicker, 1950) and trimethylamine N-oxide (Wang et al., 2019). Using this setup, it was possible to visualize and quantify the three substances p-cresol, p-cresyl sulfate and phenol out of aqueous solutions.

**Table 1.** Experimental parameters of the used MCC-IMS.

|  |  |
| --- | --- |
| Preseparation | Multicapillary chromatographic column OV-5, 20 cm |
| Column temperature | 40 °C constant |
| Ionisation source | β-radiation ($^{63}\text{Ni}$), 550 MBq |
| Drift distance | 12 cm |
| Electric field strength | 330 V/cm |
| Shutter opening time | 300 $\mu\text{s}$ |
| Drift and carrier gas | Synthetic air |
| Drift gas flow | 100 ml/min |
| Carrier gas flow | 150 ml/min |
| Temperature | Ambient |
| Pressure | Ambient |

Only negative ionization mode is used, as phenol peaks only appear in negative mode. Hence, these substances were to analyze under physiological conditions.

Breath analysis data was studied using BB_IMSAnalyse software (ISAS, Dortmund, Germany). Peak positions for the substances of interest were known from the testing above aqueous solutions and peak intensities were compared using the region comparison tool provided by the software. The evolution of the peak intensities before and after dialysis was compared to the values of control group, as well as patients’ blood results for phenol, creatinine and urea.

After ethics approval (process number 4/12/15, Ethic Committee of the University Medical Center Goettingen) a group of six spontaneous breathing patients gave informed consent about the study. Patients with CKD, that are undergoing hemodialysis three times a week were analyzed as a patient group for this pilot study (*P01* to *P06*). Additionally, six healthy volunteers were analyzed as control (*G01* to *G06*).

Exhaled air was sampled and blood was drawn before and after hemodialysis. The exhaled air was collected using flow controlled sampling over one minute und directly analyzed using the MCC-IMS as a point of-care system (Vautz et al., 2010). The patients’ blood was tested for creatinine and urea in routine analysis and for phenol and p-cresol, using ultra performance liquid chromatography with a fluorescence detector (UPLC-FLD) (King et al., 2019). P-cresyl sulfate was determined using LC-MS/MS (Cuoghi et al., 2012). In the control group, breath analysis was performed using the same method as for the patient group, but due to the absence of treatment only done once per volunteer. Control blood samples were congruently analyzed via UPLC-FLD, but not tested for creatinine and urea. As only phenol could be detected in significant concentrations in patients’ breath samples, only the development of the concentration of this substance was investigated further.

The collected data is presented through descriptive statistics as median (minimum-maximum).

## Results

CKD-Patients were aged between 57 and 80 years (two female, four male) and all of them were non-smokers. Four of them (*P01, P02, P04, P06*) had previously undergone nephrectomy and did not have any residual diuresis. *P03* had a residual diuresis of 300 mL per day and *P05* had a residual diuresis of 1800 mL.

Phenol was detected in all six control persons’ breath analysis.

In patients *P01* to *P04*, phenol was found in the MCC-IMS breath analysis before dialysis in concentrations above limit of detection. There was no phenol signal visible in patient *P05* und *P06*.

After dialysis, phenol was detected in three patients. Following calculations were performed with the data of the phenol-positive patients (*P01* to *P04*), since here phenol was concentrated above limits of detection.

**Figure 1** shows two chromatograms from patient analysis. Shown on the left is the chromatogram for *P01* at the start of HD and on the right, the chromatogram after HD. In the first chromatogram, peaks for phenol monomer and for phenol-dimer can be seen indicating a high concentration, while the second chromatogram shows a small peak for the monomer only.

**Figure 1.**
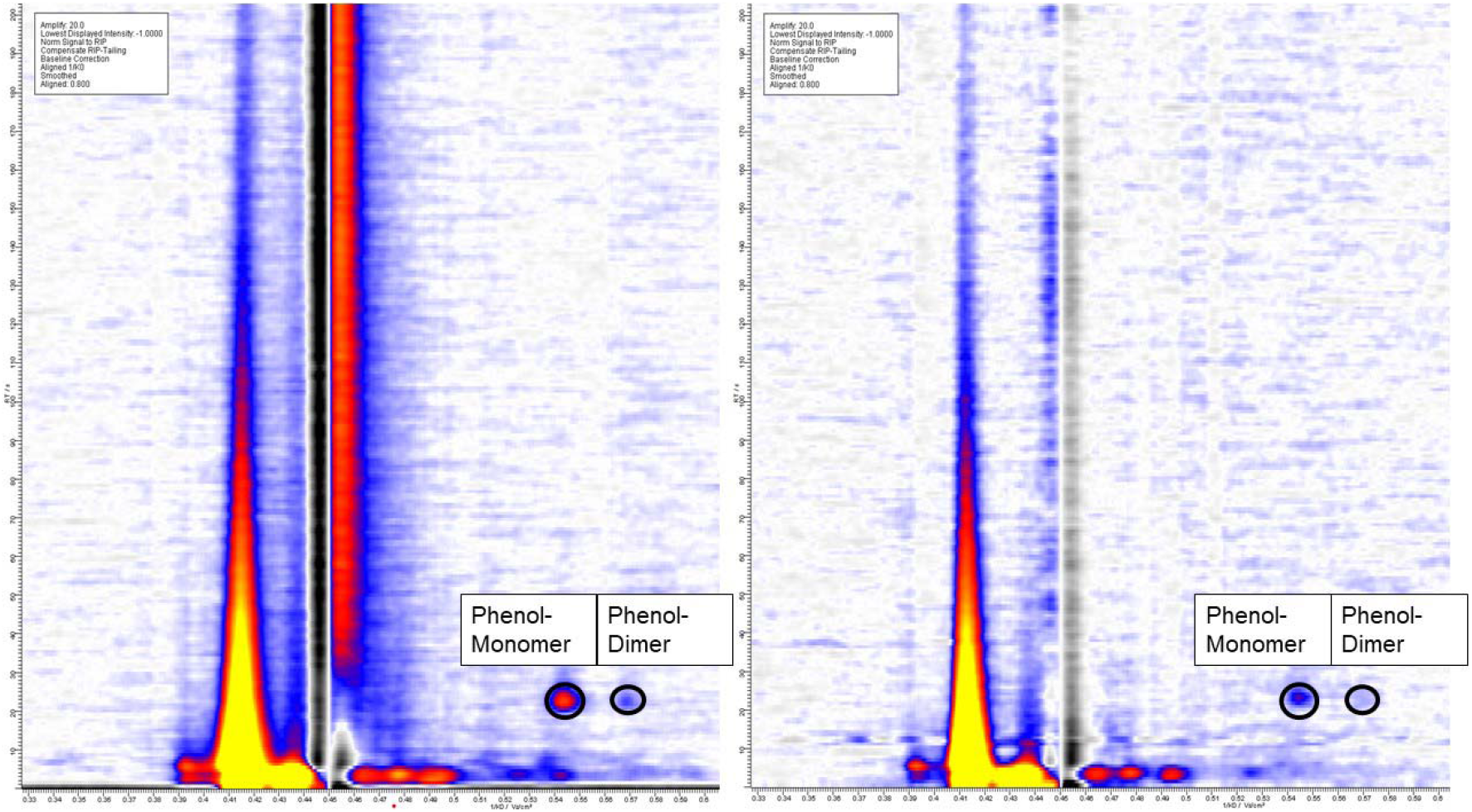
Example chromatograms from patients’ breath. Left side shows the chromatogram for *P01* before HD with peaks for phenol-monomer and phenol-dimer. Right side shows the chromatogram for *P01* after HD with a small phenol-monomer-peak and no dimer-peak.

**Figure 2** shows the development of the phenol-monomer peak-intensity for *P01* to *P04* at the start of/before HD (SD) and at the end of/after HD (ED). The signal intensity of the phenol-monomer peak was reduced from an average of 16.58 (5.42-27.28) a.U. to 7.03 (0.00-13.65) a.U.. Additionally, the measured values for the control persons *G01* to *G06* are added as control with an average of 8.50 (5.00-12.00) a.U.. The intensity of phenol monomer-peaks in patient *P03* increase from 5.42 a.U. to 5.99 a.U., all other patients showed a significant reduction of phenol in exhaled air. In total, the phenol-monomer-intensity in the patients’ breath was reduced by an average of 42.51 (−10.55-100.00) %. All patients’ peak intensities at the end of HD reached the normal range, as seen in healthy control group, with an average of 7.03 (0.00-13.65) a.U. for the patients and 8.50 (5.00-12.00) a.U. for the control group.

**Figure 2.**
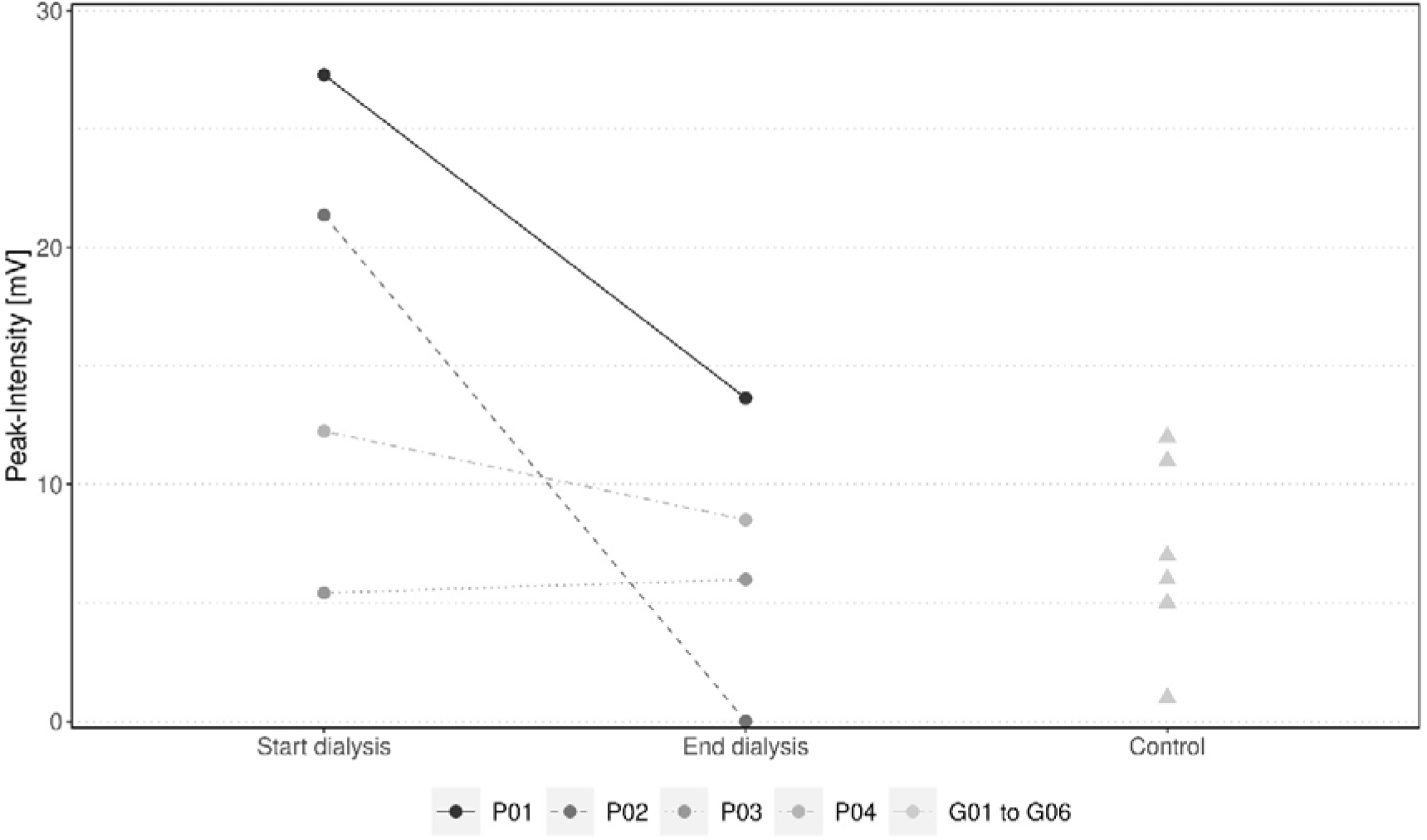
Development of the phenol-monomer peak-intensities before HD (SD) and after HD (ED) in a.U. for the patients *P01* to *P04* and control values (Control) from healthy control persons *G01* to *G06*.

All blood samples were analyzed for phenol by UPLC-FLD. **Figure 3** shows reduction of initially elevated phenol blood levels in patients *P01* to *P04* before (SD) versus after (ED) dialysis, thus after dialysis approaching normal levels of control. Blood samples from patients *P01* to *P04*, which had all tested positive for phenol in breath samples, were reduced from an average of 0.99 (0.73-1.34) mg/l to 0.34 (0.26-0.43) mg/l (control group average: 0.08 mg/l), which equates to a reduction by 64.64 (56.16-69.77) %.

**Figure 3.**
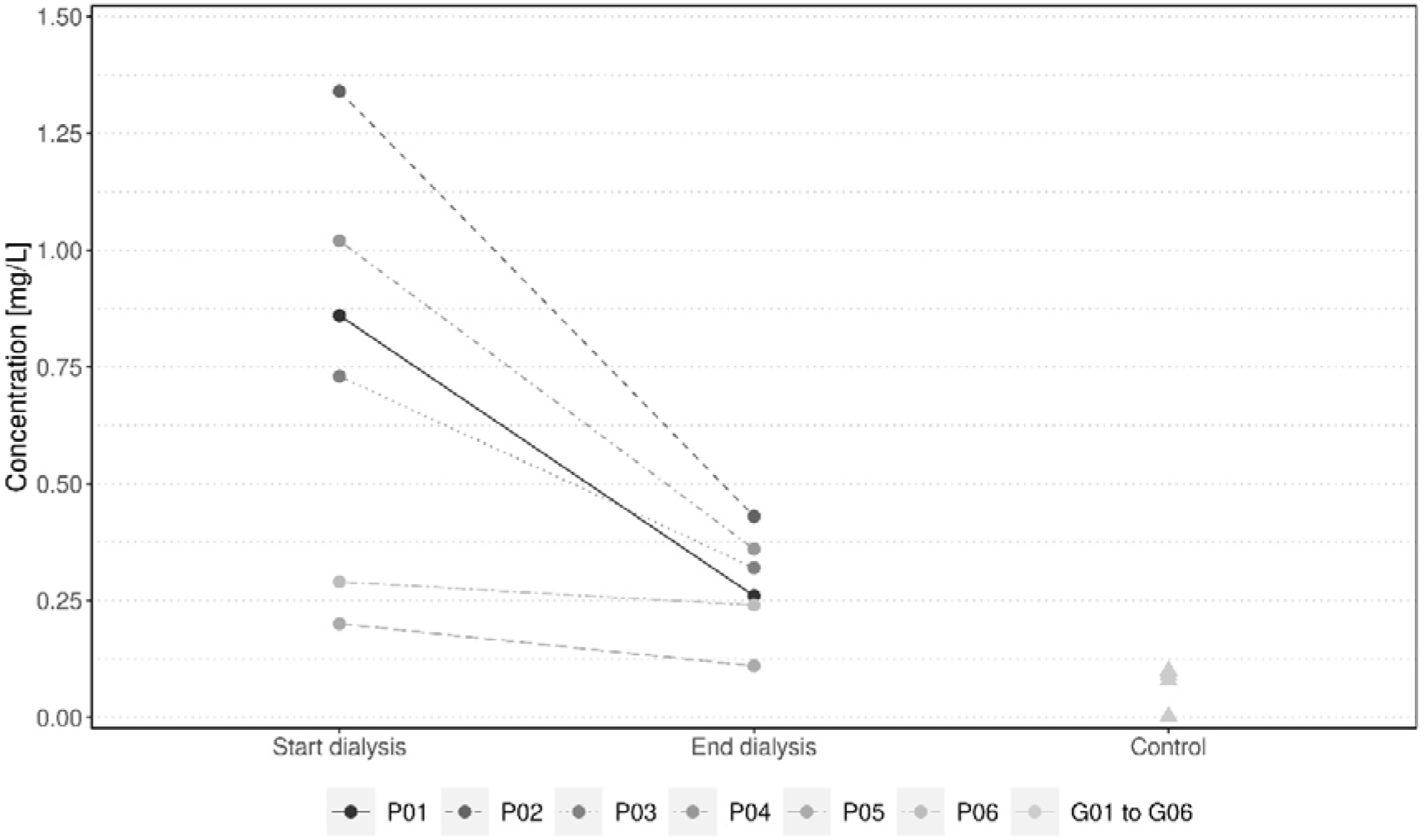
Development of the phenol-blood-concentration before HD (SD) and after HD (ED) in mg/L for the patients *P01* to *P06* and control values (Control) from healthy control persons *G01* to *G06*.

For reference, the breath-sample-phenol-negative patients’ (*P05* and *P06*) blood concentrations are shown as well. Here, reduction by 31.12 (17.24-45.00) % of phenol blood level was shown. Their absolute phenol concentrations reduced from 0.25 (0.20-0.29) mg/l to 0.18 (0.11-0.24) mg/l in average, which on its own is rather close to the range of post-dialysis-levels of phenol-positive patients or healthy individuals (control) respectively.

As a part of routine analysis, the patients’ blood was tested for creatinine and urea concentrations before and after HD. These parameters, as to be expected, mirror the trend of phenol in the blood samples, as well as breath samples (with the exception of *P03*). Creatinine concentration was reduced by an average of 63.53 (59.20-74.63) % from 11.13 (6.82-14.73) mg/dl to 4.22 (1.73 6.01) mg/dl for patients *P01* to *P04*. Urea concentration for these four patients was reduced by 70.54 (59.34-80.95) % from 65.00 (42.00-91.00) mg/dl to 20.50 (8.00-37.00) mg/dl.

Derived from our phenol peak intensities in breath analysis before HD we found that our patient collective could be divided into three groups of two (high phenol level, low phenol level, no phenol detection). Highest peak intensities were seen in patients *P01* and *P02*. These female patients were in their sixties and nephrectomized. Exhaled phenol concentrations were significantly reduced after hemodialysis.

The second group, containing two males, with low peak intensities before HD (*P03* and *P04*), were both in their fifties and nephrectomized. Phenol concentration in *P04*’s decreased after HD, while *P03*’s increased minimally.

Two male patients, *P05* and *P06*, forming our third group showed no elevated breath-sample-phenol-levels before or after dialysis. Aged over 75 years old, both hadn’t undergone nephrectomy.

## Discussion

Our results show that phenol was detectable in breath in all healthy subjects that volunteered for control group probing, therefore breath concentrations were above our limit of detection.

Of our experimental CKD patient group four out of six patients (*P01* to *P04*) presented with detectable phenol levels in the MCC-IMS breath analysis before dialysis. The remaining two individuals showed no visible phenol signal. After dialysis three patients showed measurable phenol levels, in addition to one other with possibly so far reduced levels that they were no longer detectable with our equipment settings. Of the four detectable samples, three levels were decreased, and one level minimally increased after dialysis.

In healthy adults as in our control group, phenol peak intensities in exhaled air presented low, with an average of 8.5 a.U.. Peak intensities in patients *P01*-*P04* after HD were comparable to those of healthy individuals with an average of 7.03 a.U..

Meanwhile, with an average of 0.08 mg/l, phenol blood concentration in control subjects were slightly lower than CKD-patients’ blood concentrations after HD, with an average of 0.34 mg/l. Phenol blood concentrations in literature is described for patients under RRT with 4.4 +/- 3.9 mg/l and therefore higher compared to our observations of 0.99 (0.73-1.34) mg/l before HD (Wengle & Hellstrom, 1972). Low phenol concentration was considered to be normal and is reached in phenol-positive patients after corresponding HD-treatment.

Two of our CKD test subjects did not show any measurable phenol in breath analysis. These patients were both not nephrectomized and therefore definitely had more residual kidney function than the four phenol positive patients. Considering the aromatic, charged phenol is highly water soluble and could therefore substantially be excreted through remaining kidney capacity. Blood phenol concentrations were equally lower in these patients. In contrast we also observed a negative phenol blood value in one of our control subjects, while breath analysis showed a peak in the same subject. This could of course be due to a measurement error.

The two highest phenol peaks were observed in female subjects with nephrectomies in patient history.

Another considerable factor in explaining these observations, aside from the existence of potential residual excretion due to remaining kidney function, may be the alteration in gut microbiome in these two elderly men without visible phenol peaks in exhaled breath. It has been widely recognized in recent years not only, that phenol accumulates in CKD but also that phenol, along with multiple other molecules, derives from bacteria present in the intestine and is linked to clinical outcome (Evenepoel et al., 2009). Age, gender, or simply diet dependent differences may therefore likely result in altered phenol levels in blood and therefore in breath.

From the shown measurements, phenol-monomer peak-intensities in exhaled breath (**Figure 2**) and concentrations of phenol in corresponding blood samples before and after HD (**Figure 3**) are also mirrored in blood concentrations of urea and creatinine. This data suggests that MCC-IMS is able to detect relevant phenol levels through simple breath analysis making it an interesting option for simplified and regular monitoring of RRT, especially aromatic compounds that accumulate in CKD patients, without constant lengthy procedures and costly blood work.

In further studies with larger test groups and a wide range of age, comorbidities, nephropathies etc. differences of phenol levels in breath samples need to be identified and their causes investigated to allow a better understanding of patients eligible for this kind of monitoring. An optimization of instrumentation for quantification of phenol in breath is aspired. Apart from phenol, other VOCs relevant to the pathophysiological state of CKD-patients could be identified and, perhaps in combination could more precisely mirror the actual HD efficiency.

## Conclusion

Uremic toxins accumulate in the body of patients with chronic kidney disease and need to be removed using hemodialysis. Some small toxins are transferred from the intraluminal blood to the exhaled air. These volatile organic compounds are measurable by analyzing the breath using MCC-IMS. After choosing substances as possible biomarkers, six CKD patients undergoing hemodialysis and six control persons were analyzed. Phenol could be identified as a biomarker that is detectable in the breath under physical conditions. The peak intensity for phenol changes during dialysis and it shows a correlation to the blood concentrations. The patients can be divided into groups with different dynamics of the phenol peak, that need to be evaluated further. The breath analysis of CKD patients needs to be evaluated in further studies with more patients and results need to be confirmed by multiple measurements of each subject.

## Data Availability

All data produced in the present study are available upon reasonable request to the authors

## Acknowledgements

Johannes Wieditz, statistician at UMG, created the figures 2 and 3 for publication.

## Ethical Consideration

The authors state that the Ethic Committee of the University Medical Center Göttingen approved the research under process number 4/12/15.

